# Design Approaches for Developing Quality Checklists in Healthcare Organizations: A Scoping Review

**DOI:** 10.1101/2024.09.27.24314468

**Authors:** Elizabeth Kwong, Amy Cole, Dorothy Sippo, Fei Yu, Karthik Adapa, Christopher M. Shea, Carlton Moore, Shiva Das, Lukasz Mazur

**Affiliations:** Carolina Health Informatics Program, University of North Carolina at Chapel Hill, North Carolina, United States of America; Department of Radiology, School of Medicine, University of North Carolina at Chapel Hill, North Carolina, United States of America; School of Information and Library Science, University of North Carolina at Chapel Hill, North Carolina, United States of America; Department of Health Policy and Management, Gillings School of Public Health, University of North Carolina at Chapel Hill, North Carolina, United States of America; Division of Hospital Medicine, School of Medicine, University of North Carolina at Chapel Hill, North Carolina, United States of America; Department of Radiation Oncology, School of Medicine, University of North Carolina at Chapel Hill, North Carolina, United States of America

**Keywords:** design methods, quality checklist, safety checklist, healthcare, scoping review

## Abstract

Quality checklists have demonstrated benefits in healthcare and other high-reliability organizations, but there remains a gap in the understanding of design approaches and levels of stakeholder engagement in the development of these quality checklists. This scoping review aims to synthesize the current knowledge base regarding the use of various design approaches for developing quality checklists in healthcare. Secondary objectives are to explore theoretical frameworks, design principles, stakeholder involvement and engagement, and characteristics of the design methods used for developing quality checklists. The review followed the Preferred Reporting Items for Systematic Reviews 2020 checklist. Seven databases (PubMed, APA PsycInfo, CINAHL, Embase, Scopus, ACM Digital Library, and IEEE Xplore) were searched for studies using a comprehensive search strategy developed in collaboration with a health sciences librarian. Search terms included “checklist” and “user-centered design” and their related terms. The IAP2 Spectrum of Participation Framework was used to categorize studies by level of stakeholder engagement during data extraction. Twenty-nine studies met the inclusion criteria for this review. Twenty-three distinct design methods were identified that were predominantly non-collaborative in nature (e.g., interviews, surveys, and other methods that involved only one researcher and one participant at a given time). Analysis of the levels of stakeholder engagement revealed a gap in studies that empowered their stakeholders in the quality checklist design process. Highly effective, clear, and standardized methodology are needed for the design of quality checklists. Future work needs to explore how stakeholders can be empowered in the design process, and how different levels of stakeholder engagement might impact implementation outcomes.

## Introduction

### Quality Checklists in Healthcare

Researchers have used “quality checklist” or “safety checklist” interchangeably to refer to the safety and quality management checklists that outline current evidence-based practices, serve as mnemonic devices, focus on evaluation or performance measurement, assist in maintaining or improving the safety of an organization, or take the form of a cognitive aid or goal sheet (1). A safety and quality management checklist (hereafter referred to as “quality checklist”) is an algorithmic listing of actions to verify if an action has taken place and is used to manage and control the quality of deliverables and ensure no step is forgotten (2,3). Quality checklists have been used informally across various industries, with their widespread adoption following a notable aircraft incident in 1935. In this incident, two expert pilots were killed, and engineers on board were injured when the gust-lock was not released prior to takeoff, rendering the elevators inoperable (4,5). After the incident, Boeing developed a series of checklists for the pilots to ensure critical tasks like this were completed. Since then, checklists have become essential for aviation regulation and safety, used by pilots consistently and mandated by the Federal Aviation Administration (FAA) and other regulators internationally. This practice was subsequently adopted more broadly by the military (4,5).

In healthcare, the use of checklists was influenced by the Michigan Health and Hospital Association (MHA) Keystone Center for Patient Safety and Quality Keystone ICU project conducted between 2003 and 2005. During this project, a checklist was used to ensure adherence to evidence-based, infection-control practices, successfully reducing the risk of central line-associated bloodstream infections in intensive care unit patients (6). The development and implementation of the World Health Organization (WHO) surgical safety checklist in 2007-2008 further promoted the use of checklists in healthcare. This 19-item checklist sought to reduce medical errors and adverse events during surgery while improving the consistency of care (5,7). Subsequent healthcare research has explored the development and application of checklists in various areas, including inpatient care (8), obstetrics (9), hospital discharge (10,11), chemotherapy treatment (12), COVID-19 prevention and control (13,14), and other care areas and procedures. Checklists in healthcare have demonstrated several benefits: they serve as memory aids, ameliorating the effects of fatigue, stress, and distraction, standardize task performance, ensure adherence to best practices, and promote team communication (5,15). Notably, checklists in healthcare have also been crucial for safety management, the improvement of care processes, and the reduction of mortality and morbidity (16–18). However, despite these benefits, barriers to effective design, adoption, and implementation quality checklist design persist. These barriers include slow development and adoption, inconsistent use, and a lack of effective, standardized methodology for quality checklist development, despite research and evidence highlighting the benefits of checklists (1,19).

### Design of Quality Checklists

Although the concept of “design” has existed before the Industrial Revolution, the emergence and study of design methods originated in the 1950s and 1960s with the application of novel and “scientific” methods to problem-solving after World War II and the recognition of increased complexity in industrial products (20,21). Since then, design methods have evolved with the publication of books and articles on design methods across various industries, as well as the introduction of artificial intelligence and automation. Influential design researchers including Horst Rittel, Nigel Cross, Herbert Simon, Don Norman, and more along with design organizations such as IDEO have been credited with transforming the field and introducing concepts such as design thinking, solutions-focused problem-solving, user-centered design, and human-centered design (22). These design methods and concepts have been adopted and adapted in engineering, architecture, aviation, technology, product design, and healthcare for a variety of tools, systems, and purposes, including quality checklists.

Several studies have reported considerations, recommendations, and frameworks for the design of quality checklists, which individual organizations can adapt to their respective procedures and processes (1,23–26). These studies recognize that the content and format of quality checklists depend on their specific context, so they often provide only broad guidelines or basic methods. Nevertheless, some established methods for designing checklists include literature reviews, focus groups, Delphi consensus, task analyses, heuristic evaluation, interviews, surveys, and personal experience (19,23). Guidance for the design of quality checklists often involves a review of existing literature, consideration of the user skills, and the experience, context, systems, and environment in which the checklist is intended to be used (1,19). These guidelines advocate for the involvement of potential users and stakeholders in the design process or design team, but do not provide detailed instruction on how to engage stakeholders effectively.

Stakeholder engagement, or the process of incorporating the stakeholder in the design and development process, is being increasingly used and promoted in health research (27,28), but is incompletely described in the design and development of quality checklists in healthcare. While gathering input and insights from stakeholders has demonstrated impact, it also presents challenges, leading to literature-based recommendations on how to best engage stakeholders (25,27–30). Existing studies that have utilized varying levels of stakeholder engagement to design quality checklists acknowledge the importance of stakeholder involvement. However, more research is needed to elucidate the gaps and challenges with user engagement specifically in quality checklists design and enable researchers to develop, evaluate, and implement quality checklists more effectively in their own organizations.

While the utilization of quality checklists has been documented in various settings, there is a shortage of published guidance, rigorous design methodology, and understanding of stakeholder engagement for quality checklists, particularly in healthcare (1). Despite growing evidence and various methods for quality checklist design, a lack of a highly effective, standardized methodology for the design of quality checklists in healthcare and medicine has contributed to inconsistent use, adoption barriers, and gaps in implementation (1,5). Due to the nature and purpose of quality checklists, these issues, if not addressed, may negatively impact stakeholder satisfaction, usability, quality of care and safety. Therefore, this scoping review aims to synthesize design approaches in the development of quality checklists in healthcare organizations to better understand gaps in design methodology and stakeholder engagement that can be addressed in future studies and ultimately improve quality of care.

### Aim and Research Questions

In this review, we scope and synthesize the current state of knowledge regarding the design approaches and associated levels of stakeholder engagement for developing quality checklists in healthcare organizations. More specifically, our review addresses the following questions:

1. What are the characteristics of the design methods adopted to develop quality checklists in healthcare organizations, and how were the design methods defined and measured?
2. What theoretical frameworks and design principles were used to develop quality checklists in healthcare organizations?
3. Who is involved in the development of quality checklists in healthcare organizations, and what is the level of stakeholder engagement in the process?
4. What knowledge gaps exist in the literature in the utilization of design approaches to develop quality checklists in healthcare organizations?

In this study, a quality checklist is defined as a list of items to be performed to complete a task to verify if an action has taken place (or not), and gives information with regard to quality assurance activities and helps control the quality of deliverables (2,3). This scoping review contributes to the growing literature examining how stakeholder engagement affects the design and development of quality checklists, adds to the existing knowledge base of design approaches and frameworks, and highlights gaps and challenges for future research.

## Methods

### Data Sources and Search Strategy

A comprehensive search strategy was developed in collaboration with a health sciences librarian and executed in seven databases were included in the scoping review and searched on May 3, 2023, including PubMed, APA PsycInfo, CINAHL, Embase, Scopus, ACM Digital Library, and IEEE Xplore.

The search strategy consists of relevant search terms identified based on our research aim and research questions. For example, our search terms included “checklist,” its term variations, and related terms (e.g., “cognitive aid”, “care pathway”, “clinical pathway”, “care map”, critical path”, “clinical path”) in combination with “user-centered design” and related terms (“community-based participatory research”, “human-centered design”, “co-create”, “co-created”, “co-creation”, “co-design”, “user centered design”, “participatory design”, “design thinking”, “rapid prototyping”, “rapid design”, “participatory research”, “community-based research”, “community based research”, “action research”, “design sprint”, “agile design”, “agile method”, “agile methodology”, “agile project management”, “scrum framework”, and “agile” + “scrum”).

Following the search, retrieved citations were exported to SciWheel (reference management tool; sciwheel.com) and then uploaded into Covidence (systematic review management tool; covidence.org) for manual screening. Both SciWheel and Covidence removed duplicated citations from the aggregated search results.

### Eligibility Criteria

Eligibility criteria was developed according to the PICOT Framework (31). To be eligible for inclusion, studies had to be an empirical study involving or describing the design, development, or evaluation of a quality checklist in a healthcare organization (e.g., hospital, clinic, cancer center). Studies needed to involve a safety and quality management checklist of any format (e.g., paper or digital) as the intervention but could involve any comparator or outcome. Studies could be from any publishing year or a healthcare organization in any state, country, or location. We excluded studies that did not provide information on the design, development, or evaluation of the quality checklist intervention, as well as those that only used the checklist as part of another study or to assess another intervention. Additionally, studies published in a language other than English were excluded. Table 1 provides details on the inclusion and exclusion criteria that were used to screen search results.

**Table 1.** Inclusion and exclusion criteria.

| Selection Criteria | Inclusion | Exclusion |
| --- | --- | --- |
| Population | Study is a primary research article | Exclude studies that are not primary research articles (e.g., do not include editorials, book chapters, opinion pieces)<br>Exclude systematic / literature reviews<br>Exclude protocols |
| Intervention | Include studies that involve a safety and quality management checklist | Exclude studies that do NOT involve a safety and quality management checklist |
| Comparators | N/A | N/A |
| Outcomes | N/A | N/A |
| Timing | Studies published anytime | N/A |
| Setting | Include studies that take place in a healthcare organization<br>Include healthcare organizations in any state/country/location | Exclude studies that do NOT take place in a healthcare organization |
| Study Design | Include studies that provide or describe information on the design, development, or evaluation of a quality checklist | Exclude studies that do NOT provide or describe information on the design, development, or evaluation of a quality checklist |
|  |  | (e.g., exclude studies that just use a checklist for an experiment) |
| Language | Studies available in English (includes publications in another language, as long as an English-language version is available) | Studies in which publications are not in the English language |

### Study Selection

Two investigators (EK and AC) independently screened titles and abstracts against the inclusion and exclusion criteria. Discrepancies were resolved by a third investigator (LM). Two investigators (EK and AC) then independently screened full-text articles for inclusion, with discrepancies resolved by a third investigator (LM).

### Data Extraction and Synthesis

Data extraction was performed by one investigator (EK) by using the data extraction template provided in Covidence. After customizing the template based on our research questions, it was pilot tested with three included studies and revised to ensure all relevant data were captured. For each study, the following information was extracted: DOI, title, lead author and their contact details, year, country in which the study was conducted, healthcare organization type, study aim, study design (i.e., the framework of research methods to conduct the study), start date, end date, study funding sources, possible conflicts of interest for study authors, checklist type, theoretical frameworks or design principles used, level(s) of stakeholder engagement used, characteristics of design methods used, implementation methods or frameworks used (if applicable), population description, inclusion criteria, exclusion criteria, method of participant recruitment, total number of participants, baseline population characteristics, intervention characteristics, outcomes (e.g., mean, standard deviation, p-value), study strengths, and study limitations. Extracted data were synthesized to address each research question. Specifically, the IAP2 Spectrum of Participation framework and Vaughn’s definitions for levels of participation based on the IAP2 Spectrum of Participation framework (32,33) was used to guide data synthesis for Research Question 3 (Who is involved in the development of quality checklists in healthcare organizations, and what is the level of stakeholder engagement in the process?), classify the level of stakeholder engagement in each study, and qualify what level of participation was used by each study. Stakeholder engagement was defined as one of five levels of engagement according to this framework:

- Inform: Provide stakeholders with balanced and objective information
- Consult: Obtain feedback from stakeholders on analysis, issues, decisions, etc.
- Involve: Work with stakeholders to make sure concern and aspirations are considered and understood
- Collaborate: Partner with stakeholders in each aspect of the decision-making
- Empower: Place final decision making in the hands of the stakeholders

### Quality Assessment

While the scoping review was not focused on reported outcomes and may not have required a quality assessment, we utilized the Agency for Healthcare Research and Quality (AHRQ) Evidence-Based Practice Center (EPC) approach to interpret levels of evidence for each study (34). Using this approach, one investigator (EK) performed a quality assessment of each study within the Covidence system and a second investigator (AC) validated the quality assessment. Studies were assessed using the five EPC domains: study limitations, consistency, directness, precision, and reporting bias.

## Results

### Study Selection

The database search yielded 2551 records, of which 518 were duplicates. After excluding the duplicate articles, a total of 2033 records were identified for screening. After initial screening of article titles and abstracts, 1940 records were excluded, leaving 93 articles for full-text retrieval and review. After full-text screening, 32 articles were retained and synthesized. These 32 articles reported on 29 studies (6 articles reported on 3 studies, i.e., 2 articles were published about 1 study with different cuts of information or data presented, and were combined into 1 record, for 3 different studies). Inter-rater reliability statistics were calculated for the title and abstract review stage and the full text review stage, generating a Cohen’s Kappa of 0.632 and 0.745, respectively. The article selection and review process is detailed in the PRISMA flow diagram (35) in Fig 1.

**Fig 1.**
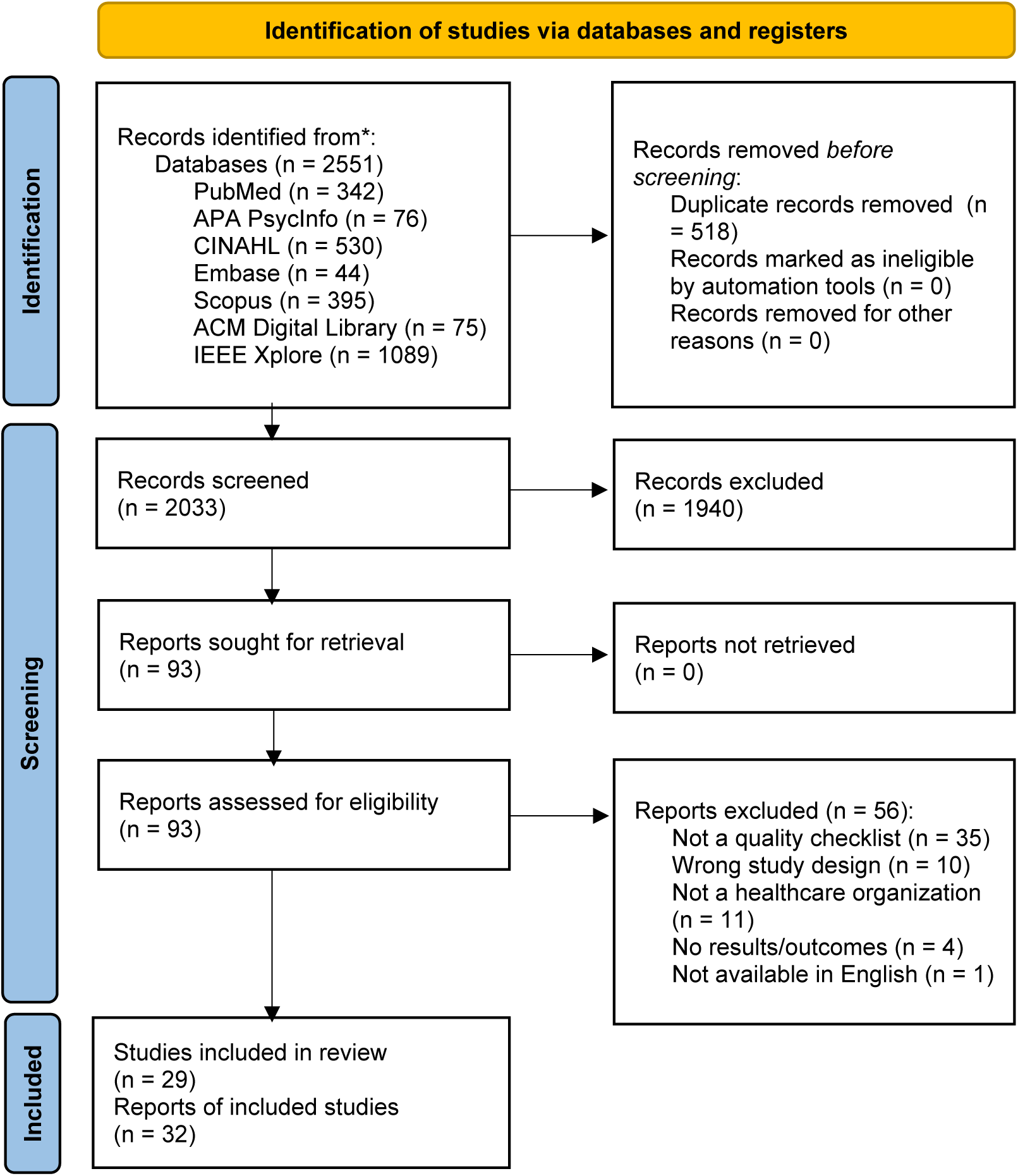
PRISMA (Preferred Reporting Items for Systematic Reviews and Meta-Analyses) flow diagram. Modified from Page *et al.*, 2021 (35).

### Study Quality Assessment

Included studies had a moderate strength of evidence overall, as they were observational in nature and not randomized controlled trials. The majority of studies (86%) were scored as “direct” (study has evidence that links interventions directly to a health outcome); remaining studies did not have measurable outcome data, intermediate data, or were only available for proxy respondents. One study (3%) (36) provided details of its power analysis, including effect size calculations and optimal information size, demonstrating a “consistent” level (same direction or similar magnitude of effect) and “precise” level (degree of certainty to reach clinically useful conclusion) for consistency and precision domains, respectively. Seventeen of the 29 studies (59%) had “low” levels of study limitations, indicating low protection against bias. These limitations predominantly resulted from small sample size and studies being conducted at one site or within one group. Reporting bias was rated as “suspected” for 66% of studies and “undetected” for 34% of studies. The “suspected” reporting bias resulted from incomplete reporting of the full study, with only a portion of the outcomes disclosed, and limitations to tested tasks and outcomes.

### Characteristics of Included Studies

Among the 29 studies identified, 34% were conducted in the United States; studies are representative of 11 different countries or regions. Studies span approximately 23 years from 2001 to 2023. Studies had between 4 and 138 participants, with a mean of 29 participants (Note: one study (37) did not provide a population number). Recruitment of participants included voluntary recruitment, convenience sampling, snowball approaches, word of mouth, social media and regular media (e.g., posters and recruitment flyers), email, and phone. Several studies cited the design, development, or evaluation of the checklist as part of a quality improvement project in the organization or system. Additionally, the majority of the study designs used were pre-and post-studies (59%).

While not a primary or secondary research question, one exploratory section of extracted evidence included documentation on whether or not the study included implementation of the checklist, and if so, what implementation methods or frameworks were included. The majority (55%) of the studies did not involve implementation while 31% of the studies discussed the implementation of the checklist, but did not describe an implementation method or framework. Additionally, 50% of the studies assessed a digital/electronic quality checklist, 12% evaluated a paper-based quality checklist, and 12% evaluated both a digital- and paper-based checklist. For additional information, see Tables 2 and 3.

**Table 2.** Overview of included studies. Summary characteristics for the 34 included studies are provided in alphabetical order by first author’s last name.

| First Author's Last Name | Year of Publication | Country | Study design | Design Methods | Theoretical frameworks / design principles | Level of stakeholder engagement | Total # of participants |
| --- | --- | --- | --- | --- | --- | --- | --- |
| Ashiru-Oredope (38) | 2022 | Tanzania, Zambia, Uganda, and Ghana (Sub-Saharan Africa) | Before and after | Modified Delphi | Modified Delphi process | Involve | 14 |
| Bartlett Ellis (39,40) | 2020, 2021 | United States | Before and after | Expert panel, use case | APA's App Evaluation Model | Consult | 7 |
| Bearsley-Smith (41) | 2008 | Australia | Before and after | Discussion / focus group | Action research | Consult | 18 |
| Benton (42) | 2020 | United States | Before and after | Literature review, interviews, survey, observations, simulated case | None described | Consult | 34 |
| Bentvelsen (43) | 2021 | The Netherlands | Interrupted time series | Contextual inquiry, questionnaires | CeHRes Roadmap, Minimum Viable Product (MVP) Strategy | Involve | 28 |
| Bergerød (44) | 2021 | Norway | Before and after | Reflective writing, consensus generating session | Consensus method based on a modified NGT; Organizing for Quality model | Involve | 20 |
| Bowie (45) | 2015 | UK | Controlled before and after | Mini-Delphi, content validity index exercise, literature and practice policy document reviews, expert steering group | Participatory design approach; SEIPS work system model; mini-Delphi | Collaborate | 18 |
| Campbell-Yeo (46) | 2021 | Canada | Before and after | Agile co-design sessions, expert consensus, interviews, focus groups | Agile, collaborative co-design process; Theoretical Domains Framework (TDF) | Collaborate | 20 |
| Carayon (36) | 2020 | United States | Controlled before and after | Scenario-based simulation, surveys | None described | Inform | 32 |
| Ceulemans (47) | 2022 | Belgium | Before and after | Semi-structured interviews, focus groups, pilot testing | Experience-based co-design approach | Consult | 15 |
| Chudleigh (48,49) | 2021, 2022 | UK | Controlled before and after | Interviews, group feedback, co-design workshops | Normalisation Process Theory (NPT); experience-based co-design | Involve | 42 |
| Flohr (50) | 2018 | Canada | Before and after | Observation, interviews, simple scenarios, mockups | Work domain analysis (WDA), participatory design approach | Involve | 15 |
| Higby (37) | 2009 | UK | Before and after | Process mapping | Action research process | Consult | Not described |
| Kerwin (51) | 2017 | United States | Before and after | Literature review, modified Delphi approach | Participatory design, modified Delphi approach | Collaborate | 14 |
| Kirkham (52) | 2021 | UK | Before and after | Delphi process, surveys | Delphi method | Involve | 30 |
| Kulp (53) | 2017 | United States | Before and after | Use case | In-the-wild co-design approach | Consult | 5 |
| Kuo (54) | 2019 | United States | Interrupted time series | Literature review, modified Delphi, interviews, think-aloud, scenario-based design/activities, focus groups | Activity Theory (AT) | Collaborate | 42 |
| Li (55) | 2014 | China | Case report | Co-design workshop, literature review | Co-design methodology based on organisational semiotics (OS) | Inform | 4 |
| Mastrianni (56) | 2023 | United States | Before and after | Literature/ case review, co-design sessions, near-live simulation sessions | In-the-wild co-design sessions | Collaborate | 8 |
| McIntosh (57) | 2017 | Australia | Cluster RCT | Simulation-based testing, interviews, usability testing | User-centered design | Consult | 40 |
| Moody (58) | 2001 | Australia | Before and after | Literature review, focus groups, questionnaires | Action research | Collaborate | 30 |
| Østergaard (59,60) | 2019, 2023 | Denmark | Controlled before and after | Participatory design workshops | Cross's research through design; participatory design (PD) | Collaborate | 85 nurses (approx.) |
| Rebic (61) | 2021 | Canada | Before and after | System Usability Scale (SUS) usability testing, interviews | User-centered design | Consult | 12 |
| Rose (62) | 2022 | Canada | Interrupted time series | Systematic review, interviews, touchpoint video, Delphi | Experience-based co-design, participatory research approach; three-round modified Delphi | Involve | 138 |
| Sarcevic (63) | 2016 | United States | Before and after | Literature review, usability testing with scenarios, focus groups, questionnaire | Iterative, user-centered approach | Consult | 4 (approx.) |
| Schild (64) | 2019 | Germany | Controlled before and after | Literature search, usability testing, questionnaires, think-aloud, brainstorming session, context interviews, personas, storyboarding | User-centered design | Consult | 43 (approx.) |
| Tarola (65) | 2018 | United States | Case report | Delphi consensus method, simulated scenarios, surveys | Three-iterative modified Delphi consensus method | Consult | 9 |
| Tyler (66) | 2021 | UK | Before and after | Literature review, co-design workshop, interviews, observations | Co-design approach | Collaborate | 40 (approx.) |
| Wu (67) | 2014 | United States | Controlled before and after | Observations, narrative simulation | Participatory design approach | Inform | 37 |

**Table 3.** Characteristics of included studies. . Characteristics include: country of publication, study design, year published, total number of participants, and implementation methods/frameworks used (where applicable).

| <u>Characteristics of Studies</u> | **Number of Studies** | **Percentage** |
| --- | --- | --- |
| **Country** |  |  |
| United States | 10 | 34% |
| UK | 5 | 17% |
| Canada | 4 | 14% |
| Australia | 3 | 10% |
| China | 1 | 3% |
| Belgium | 1 | 3% |
| Denmark | 1 | 3% |
| Germany | 1 | 3% |
| Norway | 1 | 3% |
| The Netherlands | 1 | 3% |
| Tanzania, Zambia, Uganda, and Ghana (Sub-Saharan Africa) | 1 | 3% |
| **Study Design** |  |  |
| Before and after study | 17 | 59% |
| Controlled before and after | 6 | 21% |
| Interrupted time series | 3 | 10% |
| Case report | 2 | 7% |
| Cluster RCT | 1 | 3% |
| **Year Published** |  |  |
| 2000-2009 | 3 | 12% |
| 2010-2019 | 11 | 44% |
| 2020-2023 | 15 | 44% |
| **Total number of participants** |  |  |
| 1-9 participants | 6 | 21% |
| 10-19 participants | 7 | 24% |
| 20-29 participants | 3 | 10% |
| 30-39 participants | 5 | 17% |
| 40-49 participants | 5 | 17% |
| 50+ participants | 2 | 7% |
| Not provided | 1 | 3% |
| **Implementation Methods / Frameworks** |  |  |
| Not applicable (did not involve implementation) | 16 | 55% |
| Involved implementation of the checklist, but did not describe an implementation framework | 9 | 31% |
| Action Research Cycle / Model of Implementation of Action Research | 1 | 3% |
| Behavior Change Wheel (BCW) and Theoretical Domains Framework (TDF) | 1 | 3% |
| Integrated pathway management approach | 1 | 3% |
| Normalization Process Theory (NPT) | 1 | 3% |
| **Checklist Type** |  |  |
| Digital / Electronic | 17 | 50% |
| Paper-based | 4 | 12% |
| Both digital and paper-based | 4 | 12% |
| Not described / discussed | 4 | 12% |

### Design Methods

Twenty-three distinct design methods were identified across the 29 studies, with many studies often employing more than one design method. Of these 23 design methods, the most common ones were interviews (41%), literature reviews (34%), simulated sessions/scenarios (31%), Delphi consensus (24%), and co-design / participatory design / consensus generating workshops (24%), which were a mix of collaborative (involve the cooperation of multiple parties working together) and non-collaborative approaches (involve one researcher and one stakeholder at a given time) (Table 4).

**Table 4.** Overview of design methods used.

| Specific Design Methods | Description / Summary | Number of Studies | Percentage |
| --- | --- | --- | --- |
| Interviews<sup>^</sup> | Meeting with users to ask questions and get information | 12 | 41% |
| Literature review<sup>^</sup> | Overview/summary of published works in a specific field of study | 10 | 34% |
| Simulation sessions / simulated scenarios<sup>^</sup> | Technique to replace and amplify experiences with guided ones that replicate aspects of real world settings (68) | 9 | 31% |
| Delphi consensus / modified Delphi / mini Delphi+ | Systematic process of using the collective opinion of panel members (69) | 7 | 24% |
| Co-design workshop / participatory design workshop / consensus generating workshop+ | Collaborative approach where designers work together with non-designers (70) | 7 | 24% |
| Surveys / questionnaires<sup>^</sup> | Set of questions for the purpose of gathering information through survey or study | 6 | 21% |
| Focus groups+ | Small group of people assembled to discuss, respond, and provide opinions and feedback (71) | 6 | 21% |
| Observation<sup>^</sup> | Observing participants in their natural environment to gather information | 4 | 14% |
| Usability testing (e.g., SUS)<sup>^</sup> | Testing how easy a design is to use with a group of presentative users (72) | 4 | 14% |
| Use case<sup>^</sup> | List of tasks or events to identify and organize system requirements | 2 | 7% |
| Expert panel / expert steering group+ | Group of experts and lay members who provide advice, input, and direction (71) | 2 | 7% |
| Think-aloud activities<sup>^</sup> | Verbalizing thoughts when engaging with a task | 2 | 7% |
| Contextual inquiries<sup>^</sup> | In-depth observation and interviews iusers' workplace while they work (73) | 1 | 3% |
| Narrative simulation<sup>^</sup> | Presents a consistently unfolding scenario to participants (67) | 1 | 3% |
| Pilot testing / prototype testing<sup>^</sup> | Small-scale test performed before larger-scale study | 1 | 3% |
| Content validity index exercise<sup>^</sup> | Quantify the strength of agreement on aspects of the content (45) | 1 | 3% |
| Touchpoint video<sup>^</sup> | Composite film representing all key touch points (crucial moments) in the experience (62) | 1 | 3% |
| Mockups<sup>^</sup> | Model or replica of a design/device | 1 | 3% |
| Reflective writing<sup>^</sup> | Participants reflect on articles in writing (44) | 1 | 3% |
| Process mapping<sup>^</sup> | Technique to visually map out workflows and processes | 1 | 3% |
| Brainstorming session+ | Group problem-solving technique to generate new ideas or solutions | 1 | 3% |
| Personas<sup>^</sup> | Fictional characters created to represent different user types (74) | 1 | 3% |
| Storyboarding<sup>^</sup> | "Freeze-frame" sketches showing scenarios of how people work with the system(73) | 1 | 3% |

\*some studies may use multiple design methods +predominantly collaborative method; ^predominantly non-collaborative method

Design methods were used in various ways for the design and development of checklists within the studies, including generating checklist content, providing feedback on checklist design and format, collecting data from testing, and improving the design through iterations. Studies also varied in their quality of information provided on design methods. For example, the use of a literature review ranged from an informal literature review (e.g., uncomprehensive literature search) to a full systematic review, with the level of detail in the reported information often minimal. For a list of design methods in each study, refer to Table 2.

### Theoretical Frameworks and Design Principles Used

The included studies specified the theoretical design framework, the design principles used, both, or neither. Sixteen different theoretical frameworks or design principles were identified across the 29 studies. Co-design approaches were the most commonly used design principles, with 8 studies (28%) reporting a “co-design approach” which included five sub-categories (Table 5). Seven studies (24%) used a Delphi or modified Delphi consensus technique, and six studies (21%) used what they called a “participatory design” approach. Description of the participatory design approach varied, with some studies using consensus generating workshops, expert steering groups, simulation sessions, usability testing, or a combination of these methods (Table 5).

**Table 5.**
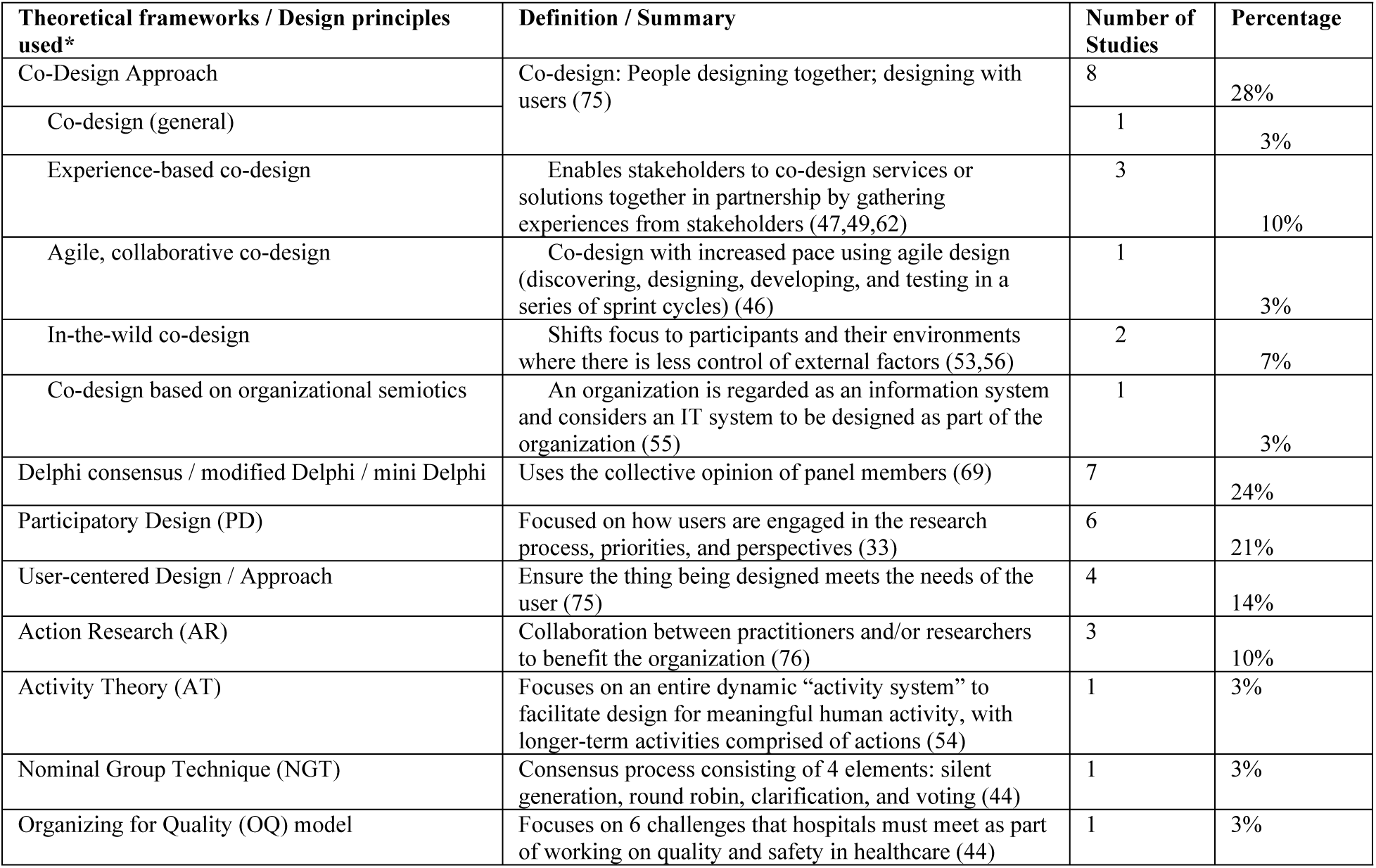
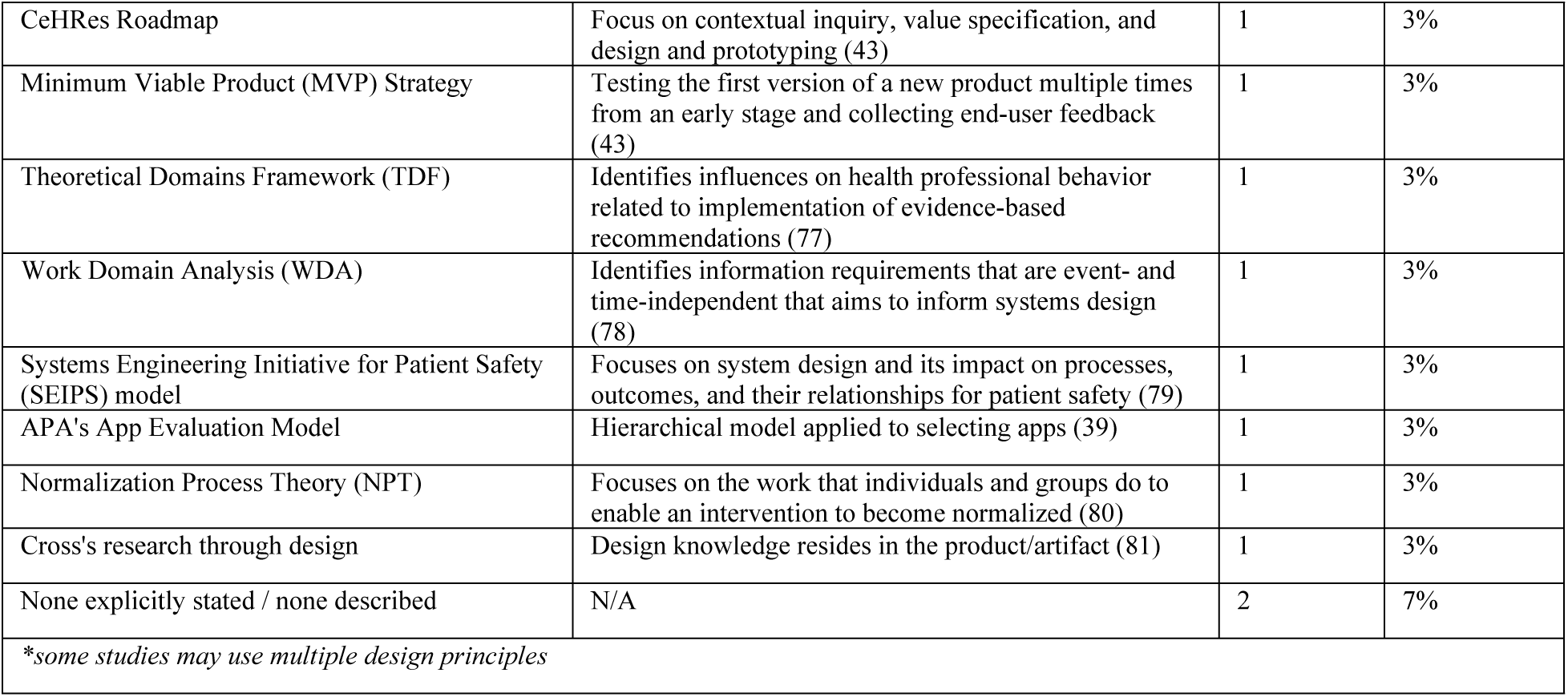
Theoretical framework and/or design principle(s) used.

### Level of Stakeholder Engagement

Studies varied in the level of stakeholder engagement according to the IAP2 Spectrum of Participation framework (Fig 2). The greatest percentage of studies were categorized as “Consult,” with 11 studies (38%) obtaining feedback from stakeholders on analysis, issues, decisions, etc. None of the 29 studies had an “Empower” level of stakeholder engagement, where final decision-making was placed in the hands of the stakeholders. The majority of studies did not explicitly state what level of participation or engagement the stakeholders had in the study.

**Fig 2.**
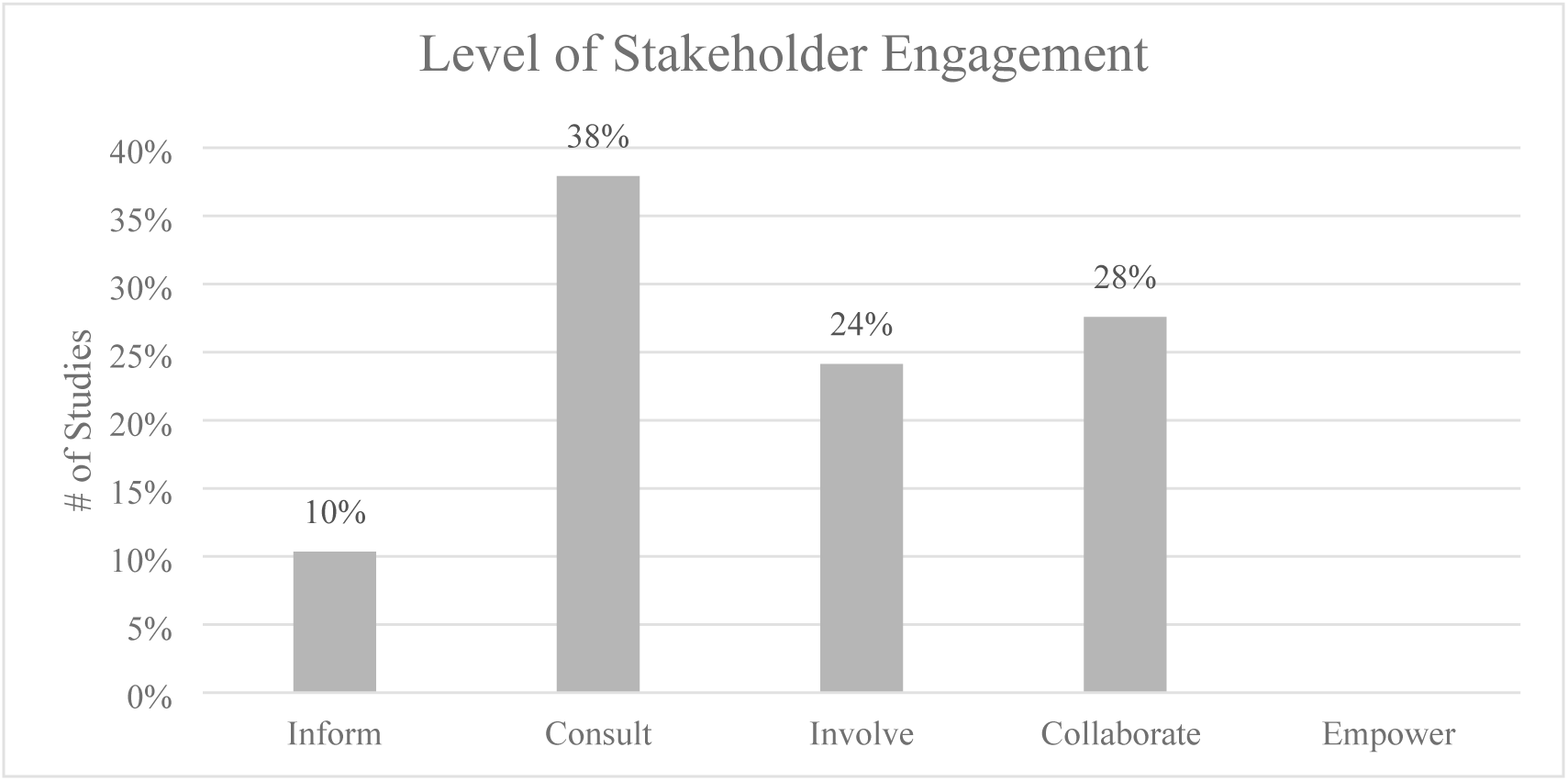
Level of stakeholder engagement. The twenty-nine studies were categorized by level of stakeholder engagement as Inform (n=3), Consult (n=11), Involve (n=7), Collaborate (n=8), and Empower (n=0).

The 9 most common design methods used among the studies were also stratified by level of stakeholder engagement using the IAP2 Spectrum of Participation framework (Fig. 3). Results demonstrate studies across the inform, consult, involve, and collaborate levels of stakeholder engagement utilized a variety of design methods, including interviews, literature reviews, simulated scenarios, and surveys. Level of stakeholder engagement was also explored in the context of funding sources available (or not available) for included studies (Fig 4). Of the 29 studies, 76% (n=22) were either nationally, organizationally, or locally/internally funded by a funding source, with 24% (n=7) listing no funding sources. Of the funded studies, the majority (n=13) were nationally funded. Of the 8 studies designated as “collaborate” levels of engagement, n=5 (63%) were nationally funded. Of the 6 studies designated as “inform” levels of engagement, n=4 (67%) did not have funding listed.

**Fig 3.**
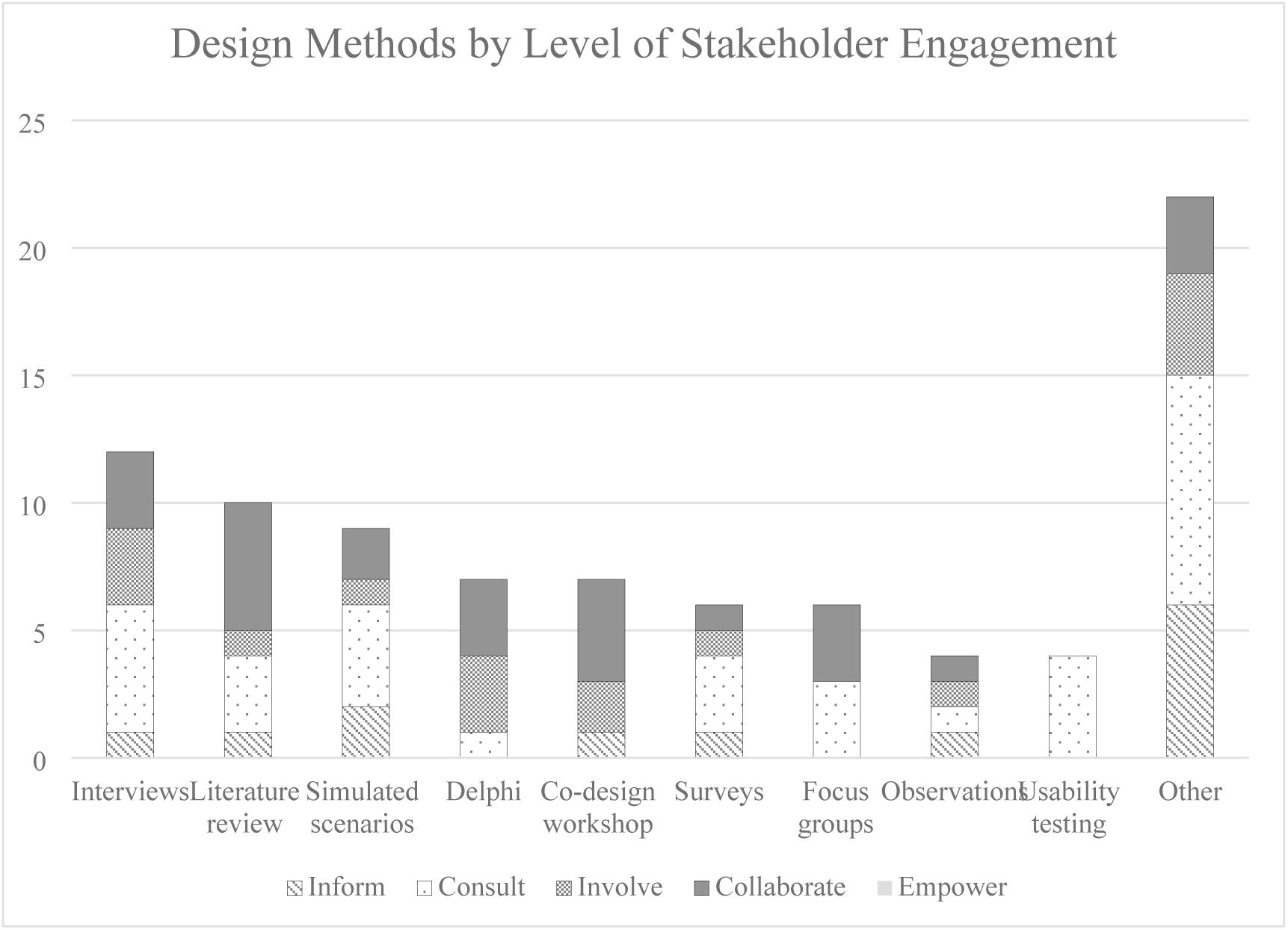
Design methods by level of stakeholder engagement. Most commonly used design methods in relation to level of stakeholder engagement (inform, consult, involve, collaborate, and empower).

**Fig 4.**
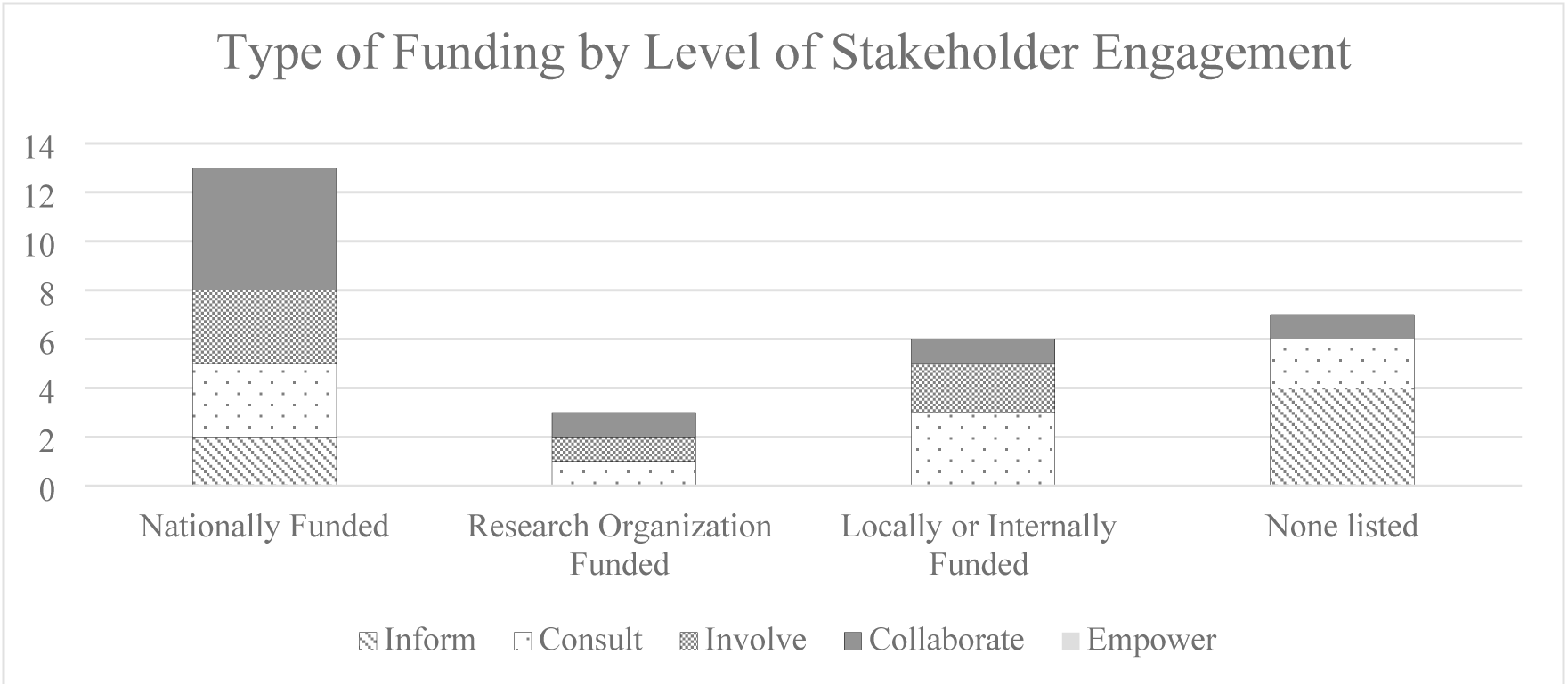
Type of funding in each study by level of stakeholder engagement. Type of funding (whether the study was nationally funded, funded by a research organization, locally/internally funded, or no description of funding at all) in relation to the level of stakeholder engagement (inform, consult, collaborate, and empower).

Stakeholder involvement, or the respective stakeholder roles/groups (e.g., nurses, physicians, patients) that participated in the study, also varied among the studies. Approximately 24% (7/29) had only one primary stakeholder group involved in the study; 76% incorporated two or more different stakeholder roles. Levels of stakeholder engagement, synthesized using the IAP2 Spectrum of Participation framework, were also stratified by one versus multiple stakeholder groups (Fig 5).

**Fig 5.**
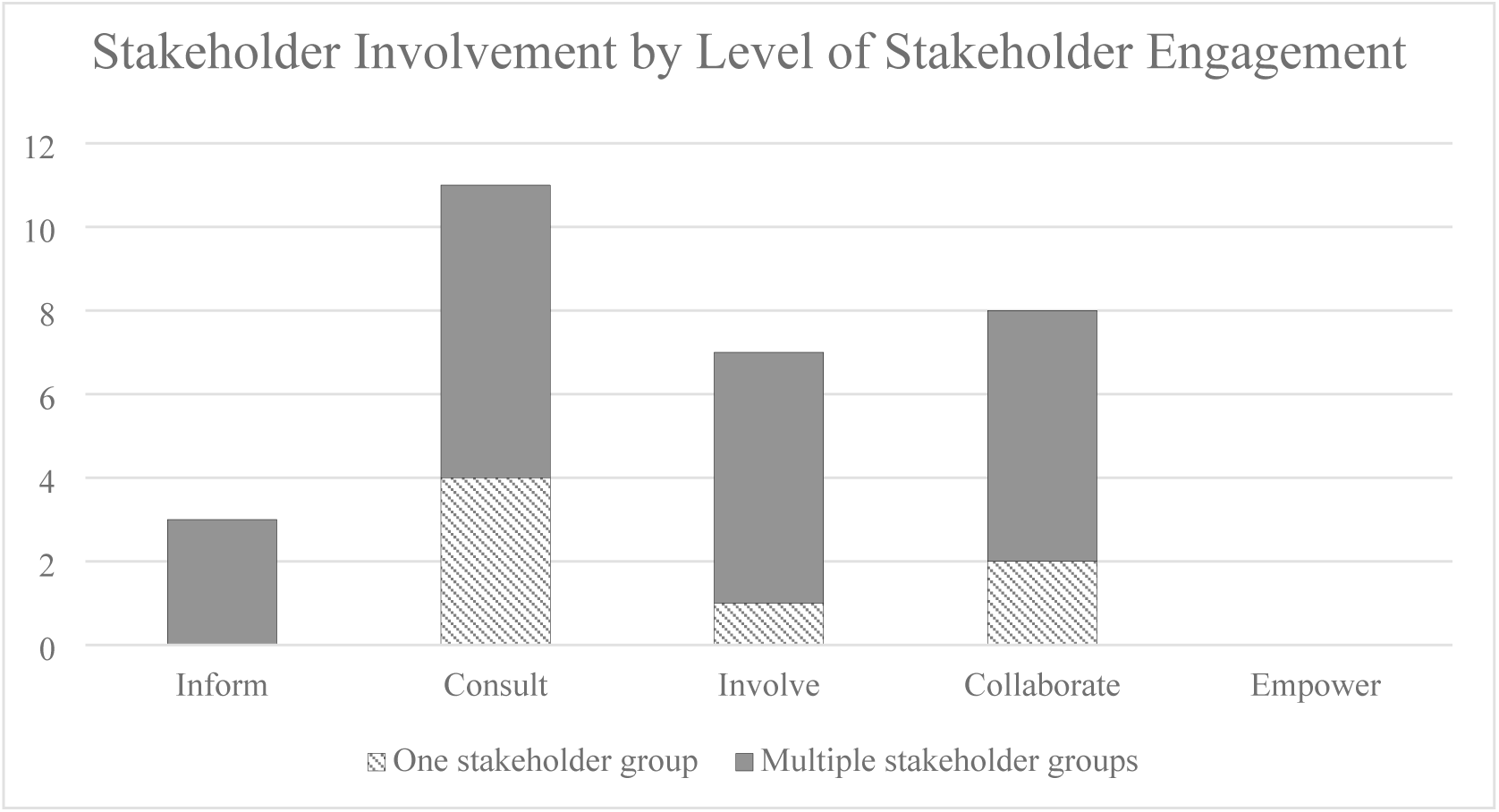
Stakeholder involvement by level of stakeholder engagement. Stakeholder involvement (one vs multiple stakeholder groups involved) in relation to level of stakeholder engagement (inform, consult, involve, collaborate, and empower).

## Discussion

This scoping review synthesized evidence from 29 studies by disclosing the current state of design approaches for developing quality checklists in healthcare organizations. We summarized major design approaches and their characteristics, adopted theoretical frameworks and design principles, and levels of stakeholder engagement.

### Design Methods

Our analysis suggests that the majority of design methods used to design quality checklists were not collaborative in nature (e.g., involve the cooperation of multiple parties working together), with most methods only involving one researcher and one stakeholder (e.g., interviews, surveys/questionnaires). Studies that used collaborative methods such as Delphi consensus, consensus workshops, and focus groups noted some logistical challenges, such as drop-out observed across consensus meeting phases, virtual-only Delphi meetings hindering productive face-to-face discussion, funding constraints, lack of speaking up in mixed groups, and limitations on participant type and locations (38,44,51,52,65). This may suggest that studies favored more feasible design methods over more collaborative yet logistically-challenging ones.

As anticipated, a wide variety of different design methods were identified among the 29 studies, and several of the studies used multiple design methods in the development of the quality checklists. A third of the studies (n=10/29) started with a form of literature review before developing their quality checklist. This trend likely stems from the diverse existing literature on checklists that can be used to inform the design and development of new quality checklists. However, several studies noted that the quality checklists discussed were part of a quality improvement project in an organization, which may arise from a local need, local evidence base, or existing system process that needs improvement, thereby not prompting a desire for a full literature review to inform the project.

Twelve of the twenty-nine studies (41%) also included a form of interviews in order to gather insights from users and stakeholders prior to checklist development and gather feedback for checklist iterations. Interviews were predominantly described as semi-structured interviews and were done with one to two research team members. All studies used interviews in conjunction with another design method (e.g., observation sessions and interviews, interviews and focus groups). This was anticipated given the growing use of mixed methods to develop a comprehensive understanding of a topic (triangulation) particularly in qualitative research (82,83). Interview methods varied by channel (video, telephone, in person), length (between 15 minutes to 1 hour), and some described using a prepared interview guide. Notably, several studies described multiple rounds of interviews for different purposes – for example, as part of contextual inquiries for data collection and for value specification, between different checklist prototypes with different focuses, or different interview purposes depending on the type of stakeholder (43,48,49,54). Across the studies using interviews, a variety of primary and occasionally secondary stakeholders were included, in line with checklist development and implementation among high reliability organizations (16). Our findings suggest that while interviews, particularly semi-structured interviews, continue to be the most widely used data collection strategy, organizations adapt interviews as a design method to fit the purposes and needs in the development of quality checklists.

Simulated sessions or scenarios were also frequently used in the design, development, and evaluation of the quality checklist (n=9/29, 31%). These studies predominantly used more than one simulated case and varied by task, environment, situation, and fidelity; for example, one study ran usability testing with multiple, simple scenarios (50), while another used near-live simulation sessions to evaluate features of the checklist (56). Given the healthcare settings of these studies, simulated sessions provide an effective way of performing testing and iterative evaluation of a quality checklist without risking harm to stakeholders such as patients in an emergency care setting (57). Some studies acknowledged that a lack of objective observations in actual settings impacted the validity and limitations of the study (47,54,64). As the focus of this scoping review was on the design and evaluation and not the implementation of quality checklists, many included studies remained in the simulated scenario phase and did not address the transition from simulation to actual (e.g., clinic) settings, although they discussed the intent to apply the checklists in real-world settings in the future.

Other design methods used (Table 4) include 11 different methods that were used in only 1 of the 29 studies. Many of these methods include modeling methods that support conceptual designs or allow the research team to visualize users and their needs, such as personas, storyboarding, and process mapping. These methods were also used in conjunction with other, more commonly used design methods. This may be because the majority of studies had a clear understanding or direct participation of checklist users to qualify data and provide feedback that make these creative modeling methods noncompulsory for research teams. These results indicate opportunities in the design methods of quality checklists that need further exploration. For example, methods such as storyboarding and touchpoint videos have been used with interviews and focus groups to expand the contextual knowledge gathered in the design of electronic patient reported outcomes (84,85). Further research is needed to examine the feasibility and potential added benefit of modeling and creative design methods in conjunction with more established design methods such interviews, focus groups, and simulated sessions/scenarios.

### Theoretical Frameworks and Design Principles Used

Studies varied in the way design frameworks and principles were reported. For example, 28% (8/29) of studies described a “co-design approach” which was further described as experience-based, agile/collaborative, in-the-wild, or based in organizational semiotics. The application of co-design methods differed across studies, involving different data gathering techniques (e.g., semi-structured interviews, focus groups, co-design sessions/workshops, use cases, literature review, simulated sessions/scenarios, Delphi consensus group, touchpoint video, and observations). This diversity was somewhat expected given the range of healthcare organization types and specific functions for each quality checklist. Similarly, studies using participatory design (n=6/29) included a variety of design methods, including consensus workshops and meetings, simulated scenarios, mockups, interviews, and questionnaires. Vaughn *et al.* describe participatory research methods and the variety in methods and tools that are participatory by design (33). Rather than emphasizing specific design methods, participatory design focuses more on how the users and stakeholders are engaged in the research process (33). Despite often nuanced differences in practice, we postulate that co-design (28%), participatory design (21%), and user-centered design (14%) approaches were among the more prevalent design approaches due to study inclusion criteria specifically looking at the design, development, and evaluation of quality checklists (Table 1).

Although not the primary focus of our review, there was a small but meaningful amount of overlap between theoretical frameworks and design principles used and implementation methods or frameworks adopted. While 55% of studies did not involve implementation of the checklist and 31% of studies involved implementation but did not describe an implementation framework (Table 3), 4 studies integrated implementation frameworks with the design and development principles (46,48,49,55,58).

These studies used frameworks such as Action Research / the Action Research Cycle, Theoretical Domains Framework (TDF), a co-design methodology for an integrated pathway management approach, and the Normalization Process Theory (NPT). Eleven of the sixteen distinct theoretical frameworks or design principles were identified in only 1 of the 29 studies (Table 5). These theoretical frameworks and design principles appear to be specific to the type of quality checklist, environment, or checklist purpose within the study. For example, one study used the American Psychiatric Association’s (APA) App Evaluation Model to support decision-making for a digital health checklist because of its application to selecting apps (40,86). Another study used the Systems Engineering Initiative for Patient Safety (SEIPS) work system model which focuses on system design and its impact on processes, outcomes, and their relationships for patient safety, to identify safety hazards for a preliminary safety checklist (45). Further research is needed to identify and evaluate the optimal use of theoretical frameworks for the development and implementation of quality checklists.

Overall, the scoping review revealed considerable variation in design quality and definition of design principles and frameworks, with no one definitive framework emerging as preferable or exceptional.

Some studies used participatory design, co-design, and/or user-centered design interchangeably or in combination when describing the study methods or design strategies (45,46,49,60,62). Other studies stated a particular approach was used but provided limited information on why the approach was used, definitions for the principle or framework mentioned, and how these principles and frameworks impacted the methods or work accomplished (37,51). Notably, the majority of studies using a participatory design, co-design, or user-centered design approach did *not* include nor clearly specify a validated theoretical framework (47,50,51,56,57,59,60,63,66,67).

### Level of Stakeholder Engagement

Level of stakeholder engagement did not appear to be associated with the design approaches or methods used (Fig. 3). For example, the 8 studies that used co-design approaches had different levels of stakeholder participation and engagement, including consult (obtain feedback on analysis, issues, decisions, etc. from stakeholders), involve (work with stakeholders to ensure concerns and aspirations are considered and understood), and collaborate (partner with stakeholders in each aspect of the decision-making) levels. However, of the design methods used in the studies categorized as “consult” levels, the majority were non-collaborative methods in nature (e.g., individualistic methods that only required one stakeholder at a given time, i.e., surveys, interviews). In comparison, methods used in “collaborate” levels of engagement studies often involved design methods that brought stakeholders altogether (i.e., co-design or participatory design sessions, Delphi consensus groups, and focus groups). This suggests a shift towards partnership with multiple stakeholders at once with increased levels of engagement. However, none of the studies were categorized as having “empower” levels of engagement (placing the final decision-making in the hands of stakeholders). This may be because studies developing a quality checklist may not have or require the level of resources needed to empower their stakeholders, or the organizations may not have the capacity to turn over full or final decision-making to stakeholders (87). Nevertheless, this absence suggests a gap in the literature, indicating empowerment of stakeholders to make final decisions in the design of quality checklists is a potential area warranting further exploration.

The level of stakeholder engagement was also synthesized in the context of funding source availability. We found a potential association of increased levels of stakeholder engagement (i.e., “collaborate” levels of stakeholder engagement) and the presence of national funding, compared to less engagement by stakeholder (i.e., “inform” levels of stakeholder engagement) with no disclosed funding (Fig 4). Specifically, 63% of studies that used “collaborate” levels of engagement (partnered with stakeholders in each aspect of the decision-making) having some type of national funding source, while 67% of studies using “inform” levels of engagement (provide stakeholders with objective information) did not cite funding of any type. This suggests studies with funding likely have more resources for stakeholder engagement and therefore are more capable of engaging stakeholders. Literature has demonstrated the importance of research funding towards meaningful stakeholder engagement and increased financial efforts and support to engage stakeholders in the design of interventions (88–90). Further research is warranted to understand how funding sources may impact design methods and stakeholder engagement in healthcare organizations.

The type of stakeholder involved in the study did not appear to be associated with stakeholder engagement (Fig 5). Studies ranged from one population type (24% of the studies; e.g., just physicians in a particular department) to a variety of stakeholder groups (76% of the studies; e.g., nurses, pharmacists, speech language therapist, physicians, respiratory therapists, dieticians, social workers, survivors, and family members) involved in one study. Studies with only one stakeholder group were spread across inform, consult, involve, and collaborate levels, as were those involving multiple stakeholder groups .

One study that identified physicians as the primary user group of the quality checklist and nurses as a secondary user group conducted usability evaluations with only physicians but acknowledged that requirements of nurses (that did not participate in usability evaluations) may differ from those that were identified using only physicians in the evaluation process (64). Other studies acknowledged that further validation of the quality checklist was needed utilizing other stakeholders or a wider range of users (39,45,63,65). Some studies also used design methods for different purposes depending on the stakeholder involved. For example, Chudleigh *et al.* held semi-structured interviews with healthcare professionals to explore intervention acceptability, feasibility, and usability and perceptions of influential factors, while they conducted semi-structured interviews with parents of newborns to ascertain experiences and perceptions of the co-designed interventions (48,49). The quantity or variety of stakeholder roles participating in the study did not appear to impact the level of engagement, suggesting the potential feasibility for an increased level of engagement while maintaining a large variety of stakeholder roles involved in the study.

### Strengths and Limitations

#### Strengths

The literature review was performed by searching 7 different databases: PubMed, APA PsycInfo, CINAHL, Embase, Scopus, ACM Digital Library, and IEEE Xplore. These databases were selected by recommendation from an expert committee as they span a diverse range of healthcare, biomedical, life sciences, behavioral and social sciences, psychology, physical sciences, health sciences, nursing and allied health, computing, technology, and engineering fields for identification of manuscripts related to healthcare organizations.

Title and abstract review and full text review were performed with two reviewers (EK and AC), with a third reviewer (LM) resolving any conflicts at each step. Inter-rater reliability statistics were calculated for the title and abstract review stage and the full text review stage, generating a Cohen’s Kappa of 0.632 and 0.745, respectively. Kappa value interpretations by Landis and Koch (1977) indicate the strength of agreement for both stages was “substantial” (91). This suggests there was a substantial degree of agreement among the independent reviewers, lending validity to literature review results.

#### Limitations

Limitations to the literature review include studies with small participant sizes and observational study designs. Thirteen of the 29 studies (45%) had sample sizes of less than twenty participants, which likely influenced the design methods and approaches that were reported. Participant size may have been influenced by the number of stakeholder groups or funding sources. Studies may have also been able to achieve data saturation through the use of various design principles, with research suggesting a range of 9-17 participants were adequate in qualitative research and usability studies suggesting approximately 15-16 participants to test the usability of a design (92,93).

There was moderate strength of evidence identified by the quality assessment as interpreted using the AHRQ EPIC approach (34). Due to the nature of the literature review, which aimed to answer the primary question of, “what is the current state of utilizing various design approaches for developing quality checklists in healthcare organizations,” studies were observational studies and not conventional randomized controlled trials. As a result, this was expected to impact the study limitations, directness, consistency, precision, and reporting bias domains when assessing for quality.

## Conclusion

This literature review demonstrated the variety of design methods, theoretical frameworks, and design principles used in the design and development of quality checklists across healthcare organizations. The studies used predominantly non-collaborative design methods (e.g., interviews, surveys/questionnaires), suggesting a preference for more feasible design methods over more collaborative methods that present logistical challenges. The review also revealed design terminology discrepancies and instances where terms were used interchangeably. The analysis of stakeholder engagement indicated a gap in studies that empowered their stakeholders in the quality checklist design process. Future research needs to clarify theoretical frameworks or design principles and justify the selection of specific design methods, and examine how they impact the outcomes. Further research is also needed into optimal ways to empower stakeholders in the design process, and how different levels of stakeholder engagement might impact design and implementation outcomes. These propositions address the need for a highly effective, standardized methodology for the design of quality checklists that may improve the use, adoption impediments, and implementation barriers that exist in healthcare.

## Author Contributions

Elizabeth Kwong and Lukasz Mazur designed and conceptualized the study. Elizabeth Kwong, Amy Cole, and Lukasz Mazur performed data curation, collection, and extraction of the data. Elizabeth Kwong conducted the formal analysis of the data. Elizabeth Kwong, Amy Cole, Dorothy Sippo, Fei Yu, Karthik Adapa, Christopher M. Shea, Carlton Moore, Shiva Das, and Lukasz Mazur interpreted the data.

Elizabeth Kwong prepared the original draft of the manuscript. All authors reviewed, edited, and approved the final manuscript.

## Registration

The scoping review protocol was not registered.

## Data Availability Statement

Data underlying the findings are fully available without restriction from the time of publication. All relevant data are within the manuscript.

## Acknowledgements and Support

This work would not have been made possible without the financial support of The National Library of Medicine (NLM) Biomedical Informatics and Data Science T-15 Fellowship (T15-LM012500). We would like to give special thanks to Jamie Lynn Conklin and Jennifer Bissram for their assistance in the scoping review and their valuable time and contributions.

